# Accurate diagnosis achieved via super-resolution whole slide images by pathologists and artificial intelligence

**DOI:** 10.1101/2024.07.05.24310022

**Authors:** Kuansong Wang, Ruijie Liu, Yushi Chen, Yin Wang, Yanhua Gao, Yanning Qiu, Maoxu Zhou, Bingqian Bai, Mingxing Zhang, Kai Sun, Hongwen Deng, Hongmei Xiao, Gang Yu

## Abstract

**Background:** Digital pathology significantly improves diagnostic efficiency and accuracy; however, pathological tissue sections are scanned at high resolutions (HR), magnified by 40 times (40X) incurring high data volume, leading to storage bottlenecks for processing large numbers of whole slide images (WSIs) for later diagnosis in clinic and hospitals.

**Method:** We propose to scan at a magnification of 5 times (5X). We developed a novel multi-scale deep learning super-resolution (SR) model that can be used to accurately computes 40X SR WSIs from the 5X WSIs.

**Results:** The required storage size for the resultant data volume of 5X WSIs is only one sixty-fourth (less than 2%) of that of 40X WSIs. For comparison, three pathologists used 40X scanned HR and 40X computed SR WSIs from the same 480 histology glass slides spanning 47 diseases (such tumors, inflammation, hyperplasia, abscess, tumor-like lesions) across 12 organ systems. The results are nearly perfectly consistent with each other, with Kappa values (HR and SR WSIs) of 0.988±0.018, 0.924±0.059, and 0.966±0.037, respectively, for the three pathologists. There were no significant differences in diagnoses of three pathologists between the HR and corresponding SR WSIs, with Area under the Curve (AUC): 0.920±0.164 vs. 0.921±0.158 (p-value=0.653), 0.931±0.128 vs. 0.943±0.121 (p-value=0.736), and 0.946±0.088 vs. 0.941±0.098 (p-value=0.198). A previously developed highly accurate colorectal cancer artificial intelligence system (AI) diagnosed 1,821 HR and 1,821 SR WSIs, with AUC values of 0.984±0.016 vs. 0.984±0.013 (p-value=0.810), again with nearly perfect matching results.

**Conclusions:** The pixel numbers of 5X WSIs is only less than 2% of that of 40X WSIs. The 40X computed SR WSIs can achieve accurate diagnosis comparable to 40X scanned HR WSIs, both by pathologists and AI. This study provides a promising solution to overcome a common storage bottleneck in digital pathology.

## Introduction

Pathological diagnosis serves as the gold standard for disease diagnosis [1]. Pathology glass slides cannot be replicated or transmitted over the network, thus affecting the efficiency of diagnosis and expert consultations [2,3]. Digital pathology has facilitated the digitization transformation of traditional pathology [4,5], substantially enhancing diagnostic efficiency and representing the direction of modern pathology development [6,7].

The integration of digital pathology and Artificial Intelligence (AI) has given rise to a novel field known as computational pathology and pathology informatics [8,9]. The development of AI in pathology (AIP) is considered as a key technology to enhance the performance of pathological diagnosis [10, 11]. AIP accelerates computer-aided analysis [12, 13], contributing to various domains such as disease classification, grading, and outcome prediction, etc. [14-22].

Nowadays, digital pathology employs a high-resolution (HR) digital scanner to meticulously capture tissue details from glass slides, generating digitized high-resolution whole slide images (HR WSIs) [23]. The file size of a 40X magnification WSI (10 gigapixels, 0.25 um/pixel) typically ranges from 5 Gigabyte (GB) to 10 GB, with the scanning process for a single slide requiring a duration of 5 minutes or more [24]. In pathology departments handling tens of thousands of annual diagnoses, digitalization of pathological tissue section requires storage spanning hundreds of thousands of gigabytes and using multiple expensive scanners [25]. Consequently, the huge expense in digitization has emerged as a significant bottleneck for the clinical application of digital pathology. It also presents challenges for the training and implementation of AIP, particularly in developing nations, where there is a scarcity of a requisite number of digitized images [26].

Our aim is to devise a cost-effective digitization approach, wherein low-resolution (LR) images, such as only those at 5X magnification scanning, are acquired and stored. A 5X image contains only 1/64th (less than 2%) of the pixels found in a 40X image, leading to a significant reduction of 5X resolution file size to 1/64 compared with its 40X counterpart. For later pathological diagnosis, 5X LR images are computationally re-generated into 20X or 40X HR images to restore the necessary image details. The process of crafting HR images from LR counterparts is known as super-resolution (SR) image generation, which was initially applied to natural images and then some medical images [27-30].

To our knowledge, we have introduced a multiscale model (MSR) for the first time that demonstrates no significant difference in diagnostic consistency between 100 paired SR and HR regions of interest (ROIs) from some typical diseases [31, 32]. Subsequent studies have further confirmed that SR generation can generate high-quality pathological images [33-35]. However, there is not yet sufficient evidence to demonstrate the overall diagnose performance of SR WSIs on a large scale, in particular, whether SR images can establish accurate AIPs.

This paper makes significant contributions in several key areas. We introduced an improved SR model designed to mitigate potential color bias issues in images from various organs within the framework of MSR [32]. Subsequently, we conducted a comprehensive WSI-based evaluation on a large scale, assessing diagnostic consistency, accuracy and time, utilizing 480 HR and 480 SR WSIs from the same slides encompassing 47 diseases (such tumors, inflammation, hyperplasia, abscess, tumor-like lesions) across 12 organ systems (including brain, lymph nodes, lungs and so on).

Furthermore we demonstrated the clinical-level accuracy that a well-developed colorectal cancer (CRC) AIP can achieve on SR WSIs, highlighting the notable potential of SR in overcoming challenges on data storage, a currently major challenge in advancing digital pathology.

## Results

Three pathologists were recruited to perform diagnostic experiment. Pathologist A has 8 years of diagnostic experience and holds the title of attending physician; B has 15 years of diagnostic experience and is an associate chief physician; C has 22 years of diagnostic experience and holds the title of chief physician. The study pipeline is shown in Figure 1.

**Figure 1.**
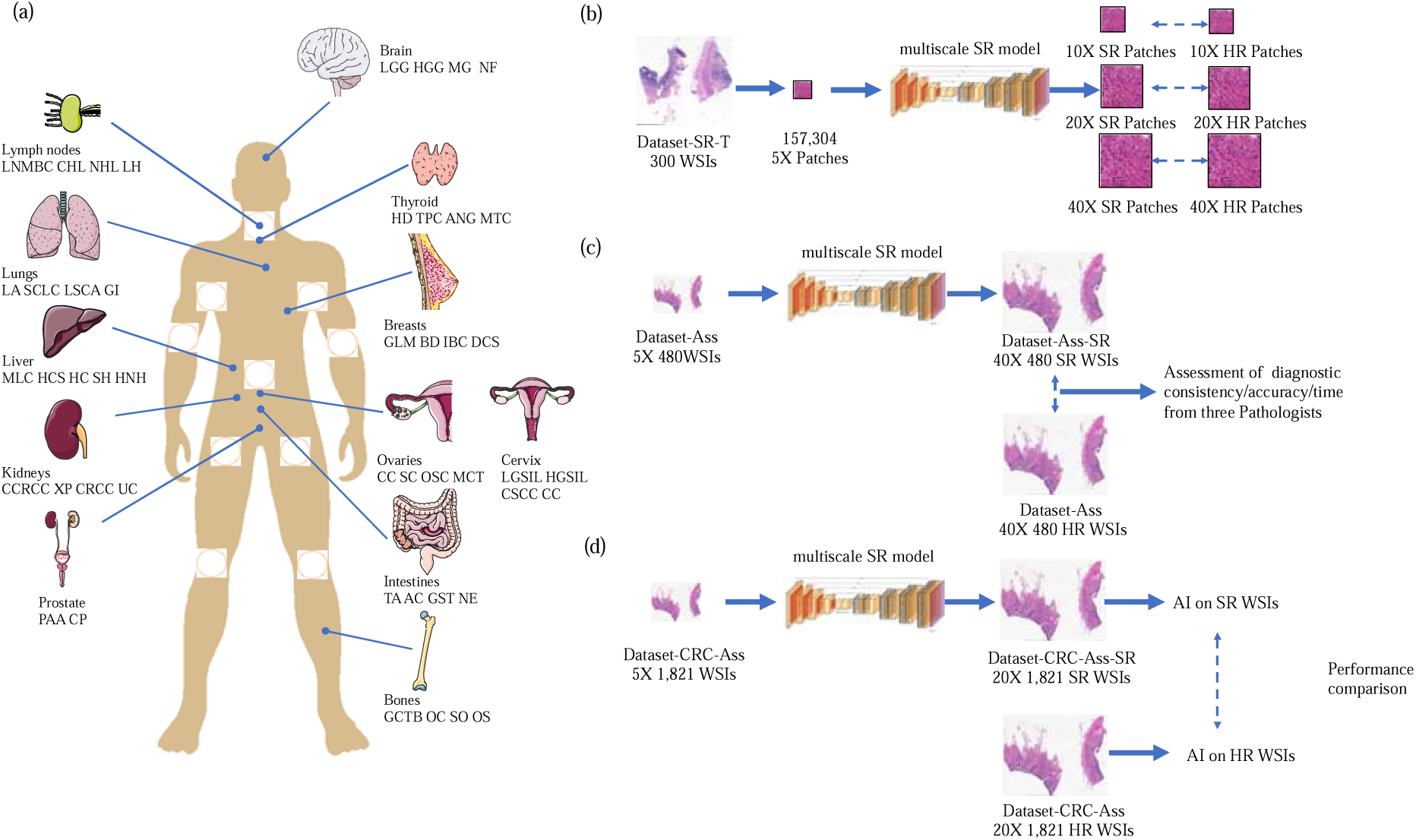
Flowchart of the study pipeline. (a) Human body parts and disease names in Dataset-Ass for clinical assessments. The brain includes four diseases: low-grade glioma (LGG, 7 subjects), high-grade glioma (HGG, 15), meningioma (MG, 15), and neurofibroma (NF, 8). Lymph nodes: lymph node metastasis of breast cancer (LNMBC, 8), classical Hodgkin’s lymphoma (CHL, 13), non-Hodgkin’s lymphoma (NHL, 17), lymphadenitis (LH, 2). Lungs: lung adenocarcinoma (LA, 15), small cell lung cancer (SCLC, 6), lung squamous cell carcinoma (LSCA, 12), granulomatous inflammation (GI, 7). Liver: metastatic liver cancer (MLC, 14), hepatic cysts (HCS, 3), hepatocellular carcinoma (HC, 16), severe hepatitis (SH, 5), hepatocellular nodular hyperplasia (HNH, 2). Kidneys: clear cell renal cell carcinoma (CCRCC, 21), xanthogranulomatous pyelonephritis (XP, 6), chromophobe renal cell carcinoma (CRCC, 7), urothelial carcinoma (UC, 6). Prostate: prostate acinar adenocarcinoma (PAA, 20), chronic prostatitis (CP, 20). Bones: giant cell tumor of bone (GCTB, 5), osteochondroma (OC, 12), suppurative osteomyelitis (SO, 10), osteosarcoma (OS, 13). Intestines: tubular adenoma (TA, 10), adenocarcinoma (AC, 16), gastrointestinal stromal tumor (GST, 6), necrotizing enterocolitis (NE, 8). Ovaries: chocolate cyst (CC, 9), serous cystadenoma (SC, 11), ovarian serous carcinoma (OSC, 9), mature cystic teratoma (MCT, 11). Cervix: low-grade squamous intraepithelial lesion (LGSIL, 9), high-grade squamous intraepithelial lesion (HGSIL, 18), cervical squamous cell carcinoma (CSCC, 7), chronic cervicitis (CC, 6). Breasts: granulomatous lobular mastitis (GLM, 8), breast diseases (BD, 9), infiltrating breast carcinoma (IBC, 17), ductal carcinoma in situ (DCS, 6). Thyroid: Hashimoto’s disease (HD, 12), thyroid papillary carcinoma (TPC, 19), adenomatous nodular goiter (ANG, 5), medullary thyroid carcinoma (MTC, 4). (b) The process involves training a multi-scale SR model using 157,304 pairs of 5X and 40X patches from Dataset-SR-T. (c) Assessment of three pathologist’s diagnostic results on paired 480 SR (Dataset-Ass) and 480 HR WSIs (Dataset-Ass-SR). (d) Comparison of patient-level diagnosis of CRC AI on paired 1,821 SR (Dataset-CRC-Ass) and 1,821 HR WSIs (Dataset-CRC-Ass-SR). The organ charts in Figure 1(a) were modified from Servier Medical Art(https://smart.servier.com/), licensed under a Creative Common Attribution 4.0 Generic License (https://creativecommons.org/licenses/by/4.0/).

We collected 47 diseases from 12 organ systems (as displayed in Figure. 1(a) legend). Each organ system comprises 40 samples, totaling 480 40X HR WSIs (Dataset-Ass, Table 1). Additionally, we gathered 300 40X WSIs (Dataset-SR-T, Table 1), divided them into 157,304 patches and downsampled 5X patches, for training the proposed multiscale super-resolution (MSR) model (Figure. 1(b)). Dataset-Ass was downsampled to 5X and then generated to 40X SR WSIs (Dataset-Ass-SR) using the MSR model, alongside Dataset-Ass, for pathological diagnostic evaluation (Figure1 (c)). To evaluate the diagnostic capability of AI on SR WSIs, we firstly trained a CRC AI (model-HR) using 20X HR patches (Table2) annotated in previous studies [22]. Subsequently, the 1,821 20X WSIs (Dataset-CRC-Ass, Table3) from the study were downsampled to 5X, and 20X SR WSIs (Dataset-CRC-Ass-SR) were generated using the proposed MSR model. The diagnostic performance of the model-HR on Dataset-CRC-Ass-SR was then compared with that on Dataset-CRC-Ass (Figure1. (d)) to evaluate whether AI trained HR WSIs can accurately diagnose the SR WSIs. More details are provided in the Methods section.

**Table 1.**
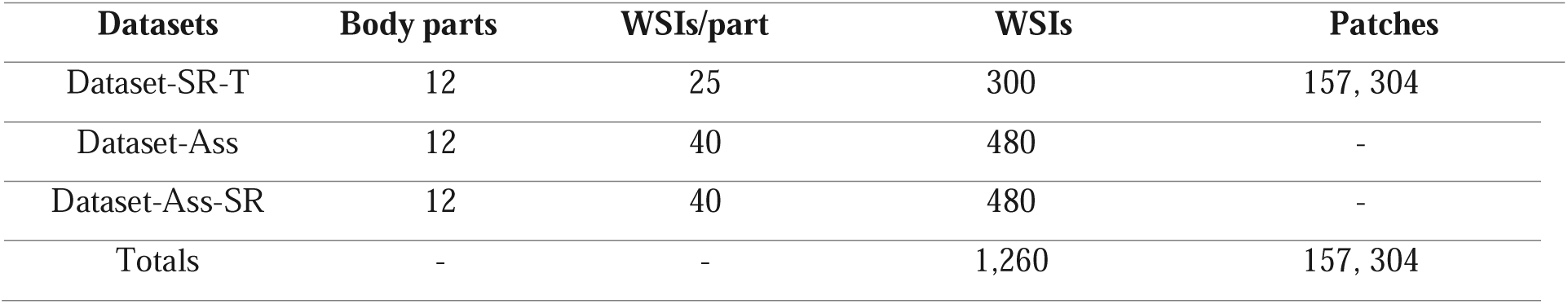
the datasets for training and clinical assessment of SR model.

**Table 2.** Patch-level training and testing dataset of CRC model.

|  | CRC |  |  | Non-CRC |  |  | Total |  |  |
| --- | --- | --- | --- | --- | --- | --- | --- | --- | --- |
|  | subjects | WSIs | patches | subjects | WSIs | patches | subjects | WSIs | patches |
| Dataset-CRC | 614 | 614 | 30,056 | 228 | 228 | 32,863 | 842 | 842 | 62,919 |
| Dataset-CRC- | 491 | 491 | - | 182 | 182 | - | 673 | 673 | - |
| Training |  |  |  |  |  |  |  |  |  |
| Dataset-CRC- | 123 | 123 | - | 46 | 46 | - | 169 | 169 | - |
| Testing |  |  |  |  |  |  |  |  |  |

-The patches in randomly selected 80% WSIs for training and the patches in remaining 20% WSIs for testing. This selection was repeated 10 times so this number varies.

**Table 3.** Patient-level diagnosis dataset for CRC model.

| Dataset | CRC |  | Non-CRC |  | Total |  |
| --- | --- | --- | --- | --- | --- | --- |
|  | subjects | WSIs | subjects | WSIs | subjects | WSIs |
| Dataset-CRC-Ass | 509 | 1,049 | 640 | 772 | 1,149 | 1,821 |
| Dataset-CRC-Ass-SR | 509 | 1,049 | 640 | 772 | 1,149 | 1,821 |

### Assessment of diagnostic consistency between HR and SR WSIs

Pathologist A-C diagnosed 480 HR WSIs (Dataset-Ass, Table1) and their corresponding SR WSIs (Dataset-Ass-SR). The Kappa test results are shown in Figure 2(a): Pathologist A (average Kappa value and standard deviation of SR and WSIs across 12 organs): 0.988±0.018; B: 0.924±0.059; C: 0.966±0.037. Except for Pathologist B in the lymphatic system (Kappa value=0.772), all other Kappa values are above 0.85 (Figure 2(b)). Figure 3 shows the frequencies of agreement correct, agreement incorrect, and disagreement of diagnosis for HR and SR WSIs. The numbers on the diagonal are much larger than those off the diagonal, indicating that consistent correct or incorrect agreements occur much more frequently than inconsistent ones. This demonstrates a high level of diagnostic consistency between HR and SR WSIs across different organs and pathologists.

**Figure 2.**
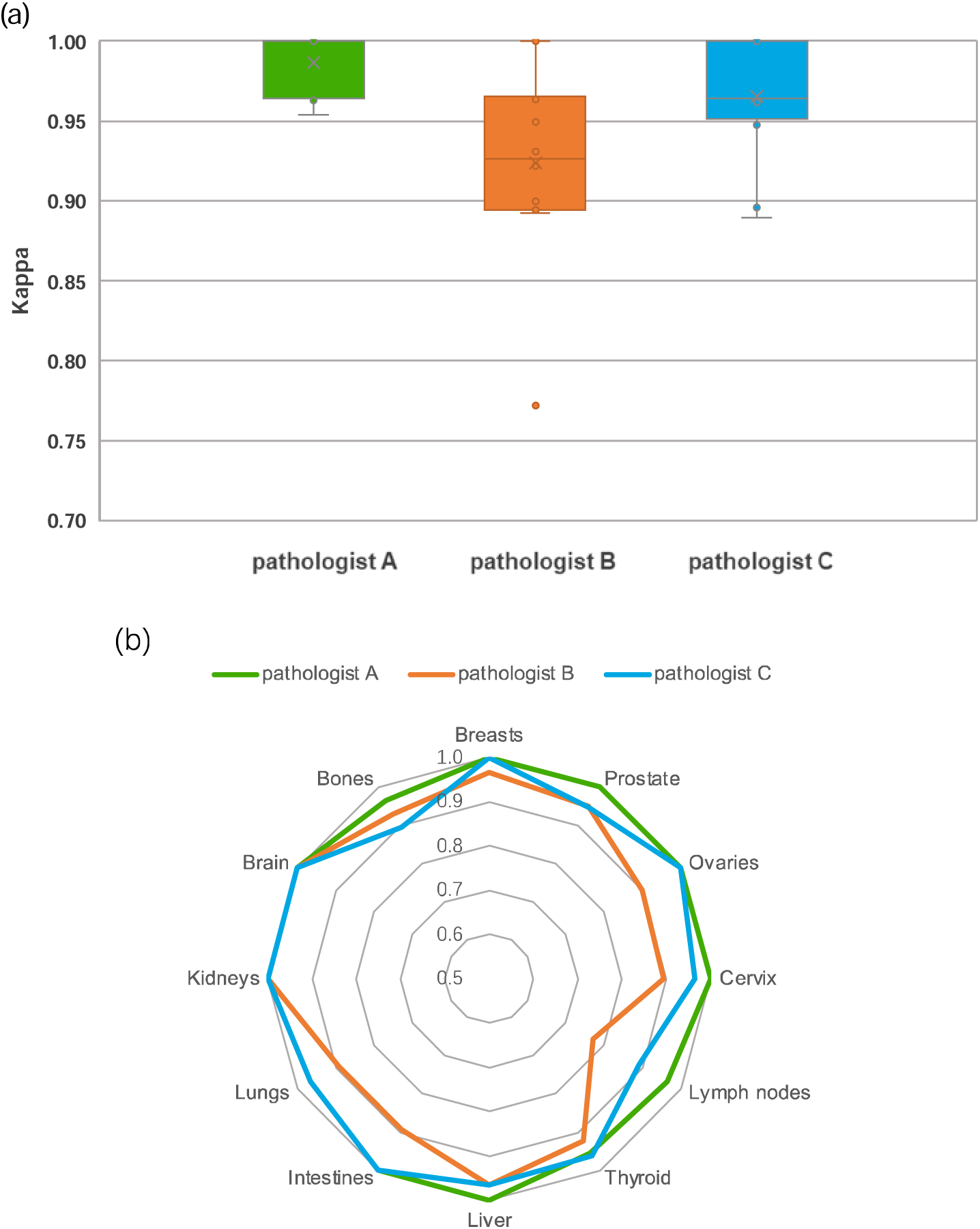
Consistency Assessment. (a) displays the distribution of Kappa values for Pathologist A-C, while the boxes indicate the upper and lower quartile values, and the whiskers indicate the minima and maxima values. The horizontal bar in the box indicates the median, while the cross indicates the mean. The circles represent data points, and the scatter dots indicate outliers. (b) lists the Kappa values for the twelve organ systems.

**Figure 3.**
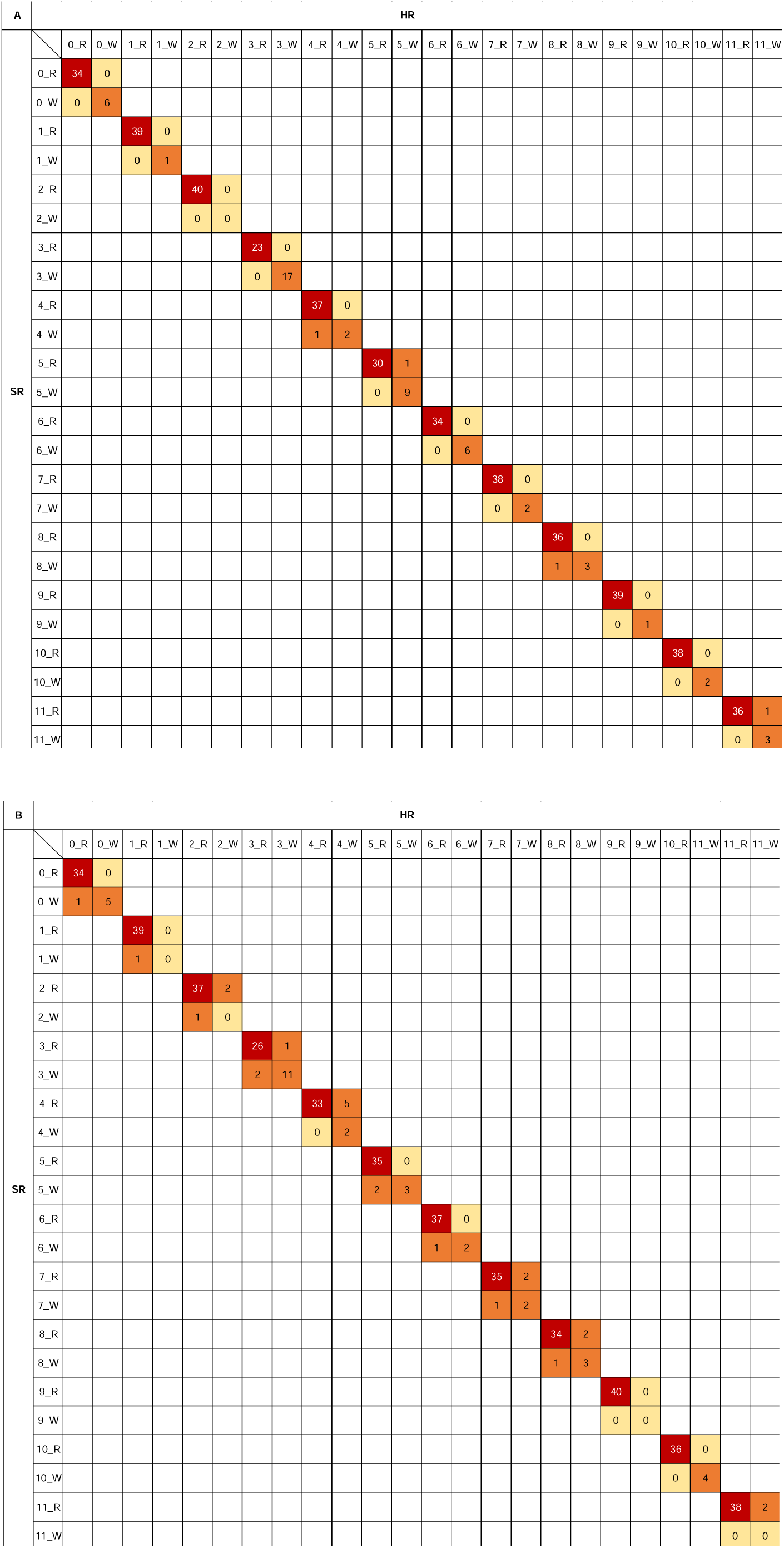
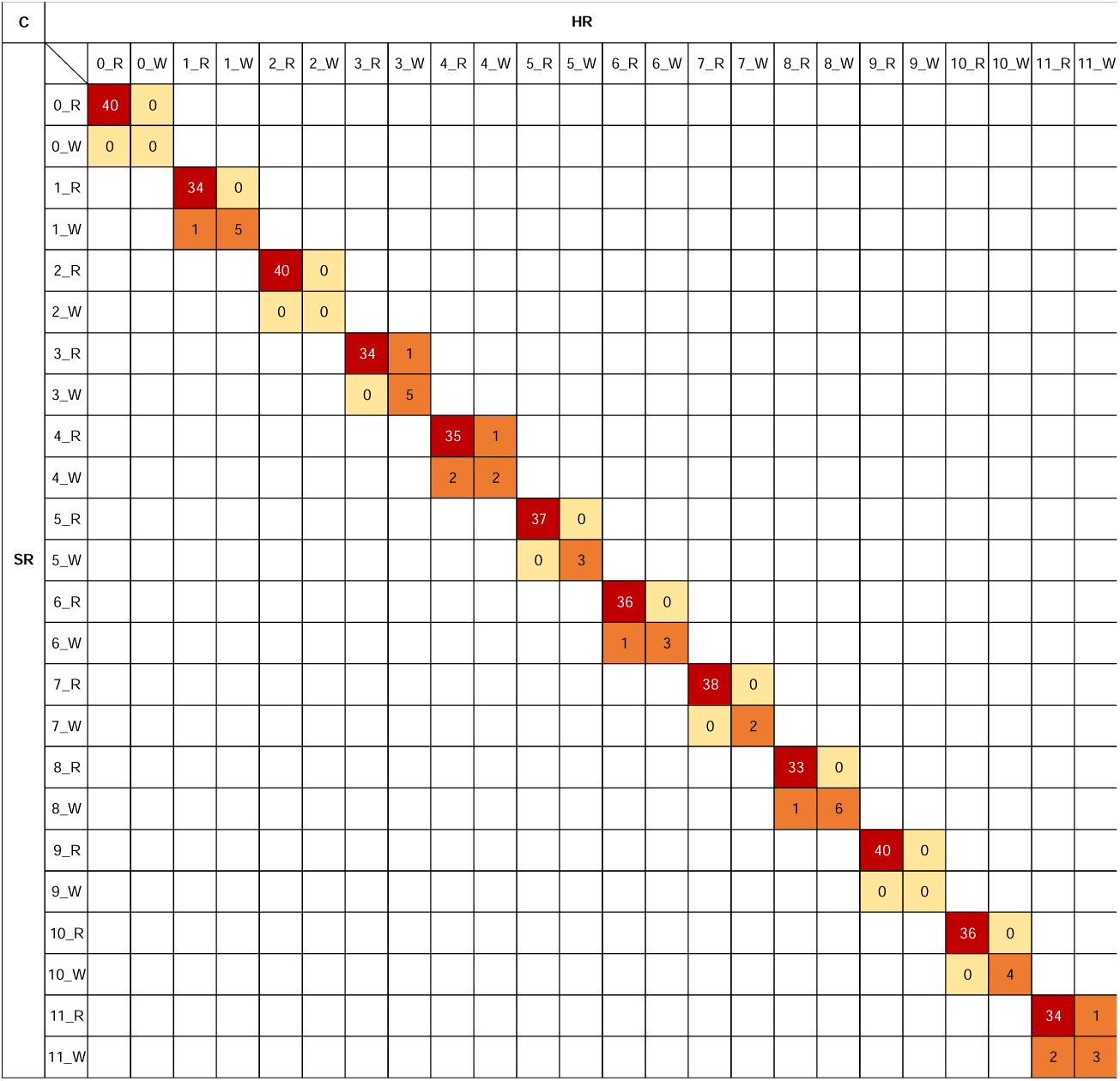
The frequencies of agreement and disagreement of diagnosis for HR and SR WSIs. The numbers in (c) represent the frequencies of agreement correct, agreement incorrect, and disagreement, R and W denote correct and incorrect diagnoses, while R and R indicate both HR WSI and SR WSI diagnoses are correct, W and W signify both are incorrect, signifying consistency. R and W denote one correct and the other incorrect, indicating inconsistency. 0-11 are the human organ systems in Figure 1(a).

### Assessment of diagnostic accuracy between HR and SR WSIs

The diagnostic results of Pathologists A-C on Dataset-Ass and Dataset-Ass-SR are compared with those of the ground truth. Figure 4(a) shows the distribution of average Area under the curve (AUC) for Pathologist A-C across 12 organ systems. Specifically, for Pathologist A, the average AUC and 95% confidence intervals on 480 HR vs. corresponding 480 SR WSIs are 0.920±0.164 vs. 0.921±0.158, with a p-value of 0.653(two-sided Paired-sample t-test); Pathologist B: 0.931±0.128 vs. 0.943±0.121, P-value=0.736; Pathologist C: 0.946±0.088 vs. 0.941±0.098, P value=0.198. Figure 4(b) displays the numerical values of the AUC for each organ in the HR and SR WSIs, where the approximate overlap of the two AUC curves indicates that the AUC values for each organ are very close. Figure 4(c) shows the sensitivity of 47 diseases by three pathologists, which doesn’t present higher sensitivity on HR WSIs than that on SR WSIs.

**Figure 4.**
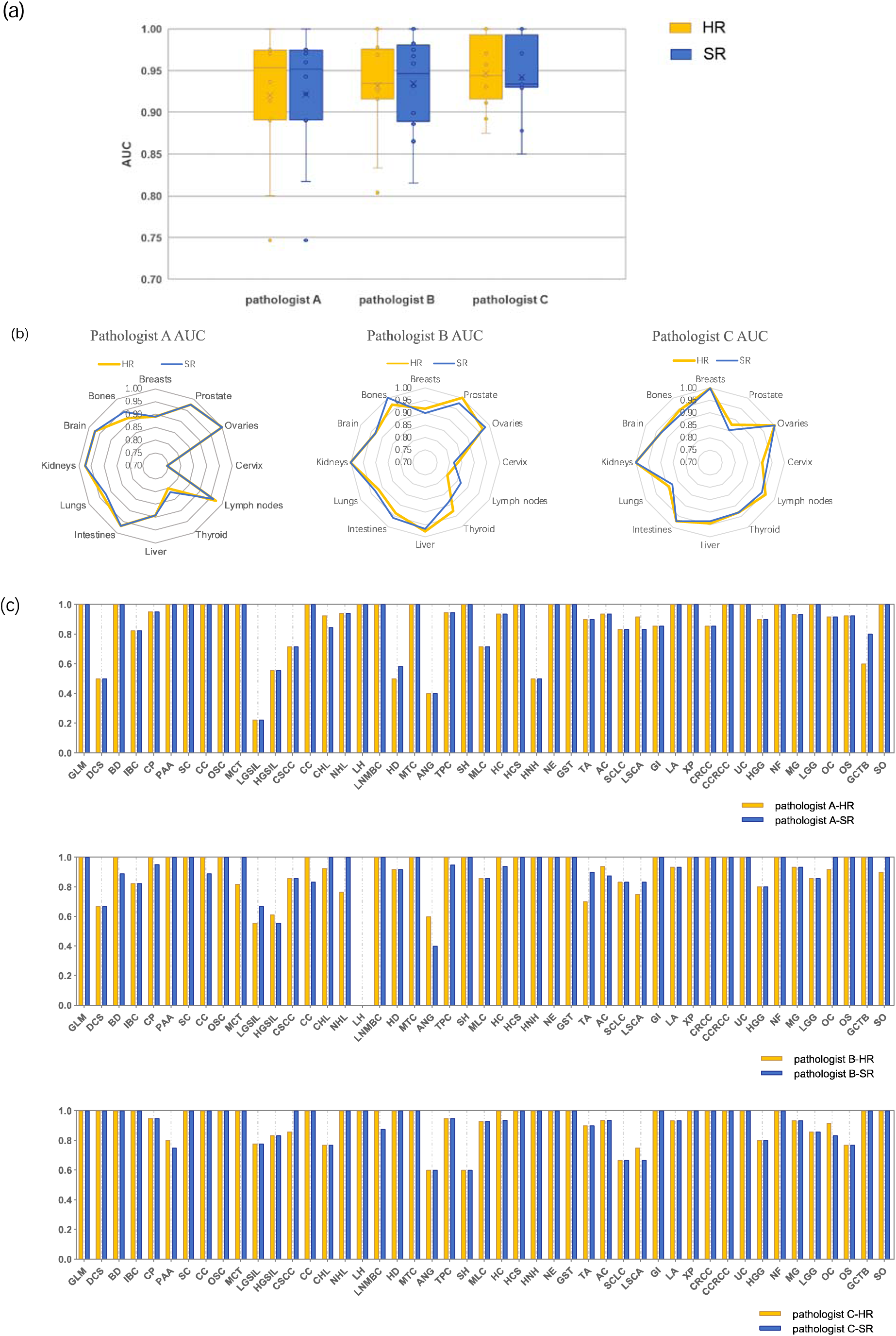
(a) Distribution of Area under the Curve (AUC) in pathologists’ diagnostic results across 12 organs. The boxes depict the upper and lower quartile values, while the whiskers extend to the minimum and maximum values. The horizontal bar within the box indicates the median, with a cross denoting the mean. Data points are represented by circles, and scatter dots indicate outliers. (b) The radar charts illustrate the names of various organs along with their corresponding AUCs. The orange lines represent the AUCs for the HR WSIs, while the blue lines represent the AUC for SR WSIs. (c) The sensitivity in 47 diseases by three pathologists.

### Assessment of diagnostic time between HR and SR WSIs

The diagnostic times for Dataset-Ass and Dataset-Ass-SR are shown in Figure 5. For pathologist A, the average diagnostic time and 95% confidence interval on HR and HR WSIs respectively are 30.877±38.7359 vs. 30.043±48.017, with a p-value of 0.489 (two-sided Paired-sample t-test). Pathologist B: 23.577±13.812 vs. 23.560±14.055, p-value=0.957. Pathologist C: 22.225±11.338 vs. 22.333±12.277, p-value= 0.768. This indicates that there is no significant difference in diagnostic time between HR and SR WSIs.

**Figure 5.**
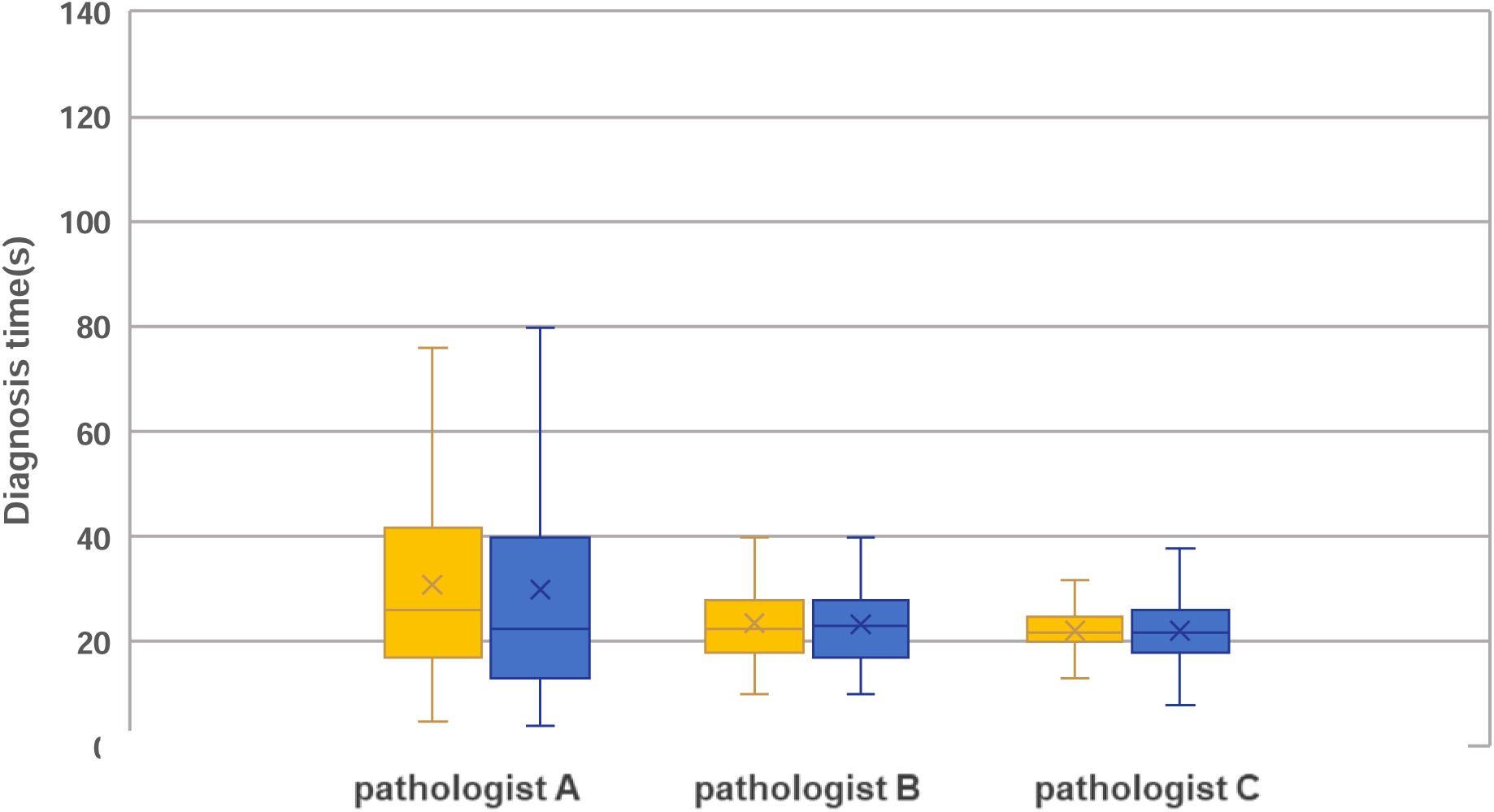
Distribution charts of diagnostic times for three pathologists on 480 SR and 480 HR WSIs across 12 organ systems. The boxes represent upper and lower quartile values, and the whiskers denote the minimum and maximum values. The red horizontal line inside the box indicates the median, while the green asterisk represents the mean.

### Performance comparison of CRC AI for HR and SR WSIs

The average AUC and 95% confidence interval of model-HR on Dataset-CRC-Ass is 0.984±0.016, but on Dataset-CRC-Ass-SR it is 0.939±0.063, showing there is a noticeable decline (P-value<0.001) on SR and HR WSIs for the CRC AI trained on HR images. This indicates that the model-HR is more sensitive to the SR images, with more significant decrease in specificity 0.901±0.131 vs. 0.971±0.035 for SR WSIs and HR WSIs. After model-HR is fine-tuned on Dataset-CRC-Training-SR, the AUC of obtained finetuned-SR (finetuned version of model-HR on SR images) on Dataset-CRC-Ass-SR is 0.984±0.013, with no significant difference compared to the AUC of model-HR on Dataset-CRC-Ass (P-value =0.810). Furthermore, there is no difference in sensitivity, specificity, and accuracy between model HR on HR WSIs and finetuned-SR on SR WSIs (Figure 6). This suggests that fine-tuning the model-HR (trained on HR WSIs) can also yield accurate AI for SR WSIs.

**Figure 6.**
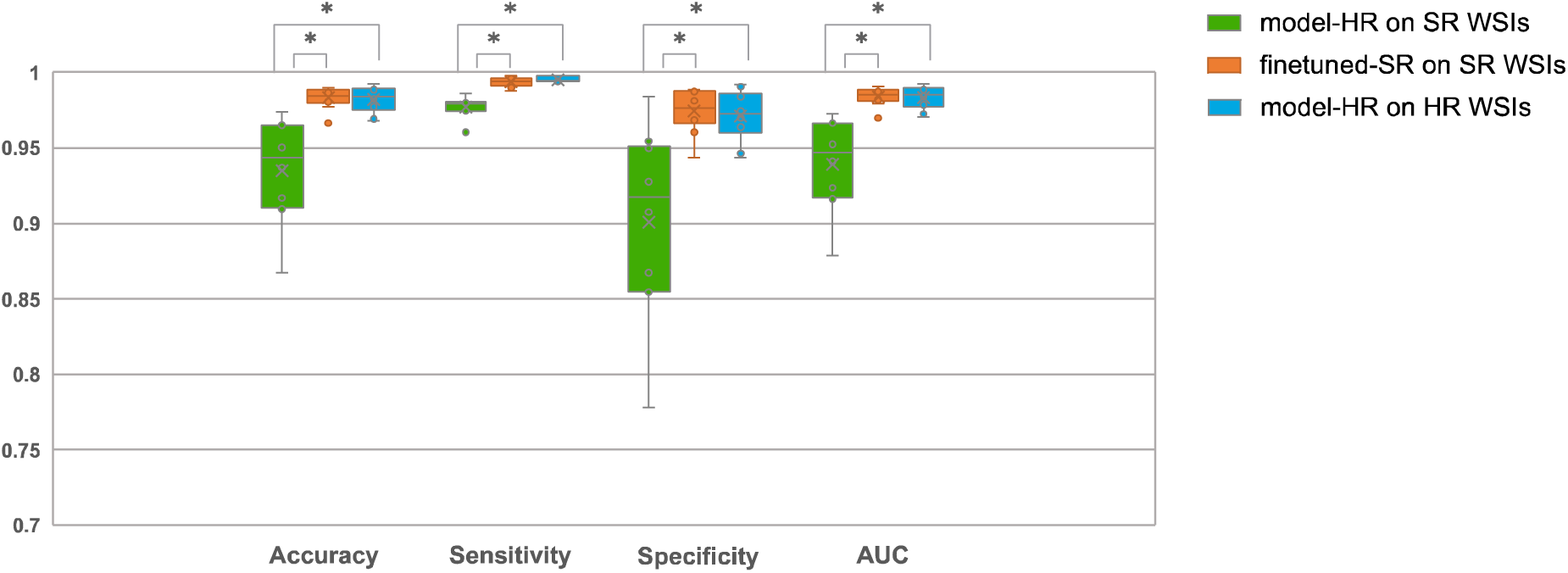
Patient-level diagnosis results on HR and SR WSIs. The average accuracy, sensitivity, specificity, AUC and 95% confidence interval of the model-HR (trained on HR images) on 1,821 HR WSIs from Dataset-CRC-Ass: 0.982±0.018, 0.996±0.004, 0.971±0.035 and 0.984±0.016; model-HR on 1,821 SR WSIs from Dataset-CRC-Ass-SR: 0.935±0.071, 0.978±0.015, 0.901±0.131 and 0.939±0.063; finetuned-SR (finetuned version of model-HR on SR images) on 1,821 SR WSIs from Dataset-CRC-Ass-SR: 0.983±0.015, 0.994±0.007, 0.975±0.031 and 0.984±0.013. The AUC of model-HR on Dataset-CRC-Ass-SR is inferior to that of model-HR on Dataset-CRC-Ass with P-value<0.001, while there are no difference between AUC of model-HR on Dataset-CRC-Ass and that of finetuned-SR on Dataset-CRC-Ass-SR with P-value=0.810 (two sided paired t-test). The boxes represent upper and lower quartile values, and the whiskers denote the minimum and maximum values. The horizontal line inside the box indicates the median, while the asterisk represents the mean. The circles represent data points, and the scatter dots indicate outliers. * denotes significant difference.

## Discussion

Digital pathology facilitates the storage, retrieval, and sharing of pathological slides, enhancing the precision, efficiency, and dependability of pathology practices. Furthermore, whole slide images (WSIs) can be analyzed through artificial intelligence, aligning with the trajectory of computational pathology’s advancement. The pathological tissue sections are scanned at high resolutions (HR), magnified by 40X or 20X is imperative to preserve these intricate image details. However, HR scanning yields sizable file dimensions that can leading to storage bottlenecks for processing large numbers of WSIs for later diagnosis in clinic and hospitals.

Theoretically, compared to 40X WSIs, 5X WSIs consist of only less than 2% pixels, leading to a 1/64 reduction in data storage requirements. Therefore, we assume that storing 5X WSIs can overcome the data bottlenecks associated with the challenge of the data storage required by the aforementioned 40X scanning. Each pixel in 5X WSIs is approximately 2 microns, allowing for the visualization of cells and structural features in tissue sections. The smallest cell for pathological diagnosis is the small lymphocytes, with a diameter of approximately 7 microns [36] which occupies 3.5×3.5 pixels in a 5X WSI. Hence, the small lymphocyte for diagnosis can be observed effectively in 5X images. Therefore, when there is obvious 5X information about the smallest structures present, it is possible to obtain a higher magnification, such as 40X, to visualize these structures in greater details by an effective method of image processing. Moreover, another benefit of 5X WSIs is the faster scanning speed, as it involves capturing and digitizing less pixel numbers compared to HR scanning.

We have demonstrated that the proposed multiscale SR method can regenerate SR WSIs that closely resemble 40X HR WSIs from 5X WSIs. Three pathologists with varying levels of experience participated in the experiment, indicating comparable diagnostic efficacy between HR and SR WSIs. Furthermore, the metrics like AUC suggest no significant differences in diagnostic accuracy. Additionally, their diagnostic times did not differ. These experiments, conducted on a large scale, confirm that the proposed method can yield accurate SR WSIs with highly consistent diagnostics with HR WSIs, providing robust support for diagnosis pipeline based on 5X WSIs and SR generation.

It is worth noting that, in order to eliminate other potential interfering factors, all experiments on diagnosis were conducted solely based on WSIs. Apart from the location of the specimens, no additional patient information from other examinations was utilized. Moreover, each organ involved 4-5 typical diseases, and each disease has samples from several to twenty patients. As a result, the AUC values for some organs experienced slight reductions, yet with the AUC values for HR and SR maintaining either consistently high or low levels without differences.

One highlight of our study is the investigation of whether the performance of artificial intelligence trained on HR WSIs can be replicated on SR WSIs. Experimental results indicate a slight decrease in performance of AI trained on HR WSIs when directly applied to SR WSIs, possibly due to the higher sensitivity of the AI than human to image details. Previous studies [46] have also noted similar variations in the performance of pathological AI across different scanners or medical centers. However, the fine-tuning on SR WSIs can achieve diagnostic performance comparable to that of HR WSIs. Therefore, SR WSIs do not lead to a decline in AI performance.

The main limitation of this study is the use of bilinear interpolation to acquire 5X WSIs, which might introduce some differences compared to real 5X WSIs from scanner directly. The primary reason is the current absence of LR such as 5X scanners. However, the deep learning can effectively learn the relations between paired images for example LR and HR images. Moreover, a significant body of SR research demonstrates that SR images can closely approximate HR images when the reconstruction conditions remain consistent with the training conditions of SR models. Therefore, the findings of this study are expected to be reproducible in real scanning systems if training and testing WSIs are from the same scanner systems. Additionally, this study was conducted solely in the histopathology images. Although this study provides the crucial evidence for the development of novel digital technologies that can overcome the existing bottlenecks in digital pathology, we believe that a broader assessment is necessary.

The next step of research involves the development of a 5X scanner. By obtaining paired 5X images on a 5X scanner and 40X images alongside a 40X scanner, the SR model of this study can be retrained to achieve the MSR model on LR scanning and SR information reconstruction for real LR scanning. This study will greatly advance the development of digital pathology by overcoming storage bottlenecks.

## Conclusions

To overcome the existing bottlenecks of huge storage requirements in digital pathology, this paper introduces an alternative approach based on LR scanning. This approach reconstructs LR WSIs into SR WSIs and undergoes extensive evaluations alongside HR WSIs, demonstrating a high level of consistency in pathological diagnosis and AI system. The findings of this study highlight the potential of a novel strategy based on LR scanning for digital pathology, which could drive advancements in the field and enhance the efficiency and accuracy of pathological diagnosis by overcoming storage bottlenecks.

## Data Availability

All data produced in the present study are available upon reasonable request to the authors

## Method

The study was conducted in accordance with the Declaration of Helsinki (2013 revision) and approved by the Institutional Review Board of School of Basic Medical Sciences, Central South University (No. 2022-KT74), and individual consent was waived for retrospective analysis.

### Data collection and review

All glass slides were prepared by Formalin-fixed paraffin-embedded tissue (FFPE) and sourced from the Department of Pathology at Xiangya Hospital, Central South University. To ensure comprehensive representation of various human tissues, the collection spans across 12 organ systems. Dataset-SR-T was utilized for MSR model training, where 25 glass slides were randomly selected from each system, resulting in a total of 300 slides. Dataset-Ass were utilized for clinical assessments, where the types of disorders were meticulously chosen to encompass malignancies, inflammatory conditions, and other illustrative disorders from various organ systems. The slides were randomly retrieved from the pathology archive by technicians by the key word including names of organ system, and then digitized utilizing a scanner (3DHISTECH Ltd., Budapest, Hungary) at 40X magnification, resulting in the acquisition of 40X WSIs, boasting a resolution of 0.12 µm per pixel.

Dataset-Ass underwent a retrospective review, accompanied by essential clinical materials, conducted by three pathologists holding the title of Associate Chief Physician and possessing a minimum of 10 years of clinical experience. Only the WSIs that met the diagnostic criteria were retained, resulting in a total of 480 WSIs available for further analysis (Table 1). The three pathologists were not participating in the subsequent research. Given that contemporary pathological scanners exclusively utilize 20X or 40X lenses, the WSIs in Dataset-Ass underwent 40X->5X down sampling using bilinear interpolation [37] and 5X->40X super-resolution generation by proposed MSR model, and the resulting 40X SR WSIs are denoted as Dataset-Ass-SR.

We also developed a CRC AI model using the data from the literature [22], which was obtained from Xiangya Hospital and scanned at 20X magnification (KF-PRO-005 scanner, KFBIO company, Ningbo City, China). The dataset, named Dataset-CRC, was created by extracting 62,919 patches of size 300×300 from 614 WSIs with cancer and 228 WSIs without cancer (Table 2).

We randomly selected 1,821 WSIs from 1,149 subjects from the literature [22] to assess the CRC model’s patient-level diagnostic capability (Dataset-CRC-Ass). These WSIs were down sampled to 5X, then reconstructed to 20X by MSR model, resulting in Dataset-CRC-Ass-SR (Table 3).

### Data preprocessing pipeline

The Dataset-SR-T were randomly divided into training and validation set comprising 240 and 60 WSIs. Each WSI was segmented into non-overlapping patches of 2048×2048 dimensions containing human tissue, and a maximum of 1000 patches were randomly selected from each WSI. As a result, the training set was formed by a total of 131,656 patches, while the validation set included 25,648 40X patches, where the paired 5X patches were subsequently resized to 256×256 dimensions using bilinear interpolation.

### Improved MSR model

We have developed a multi-scale SR approach, which occasionally leads to color discrepancies [32]. Drawing inspiration from [38], we present an improved MSR model to identify an SR image that closely matches the style of HR image within the hidden space. The model comprises three key modules: an encoder, a style transfer unit, and a decoder, as illustrated in Figure 7. The encoder transforms the input image into 20 latent vectors. Alongside the consistent input from style transfer [39], the down-sampled input images are integrated with these latent vectors using FBLOCK. This amalgamation aims to identify images that exhibit similarities to HR image styles. Subsequently, images sharing comparable styles undergo further decoding to ensure that the resulting SR images embody the authentic attributes of HR images.

**Figure 7.**
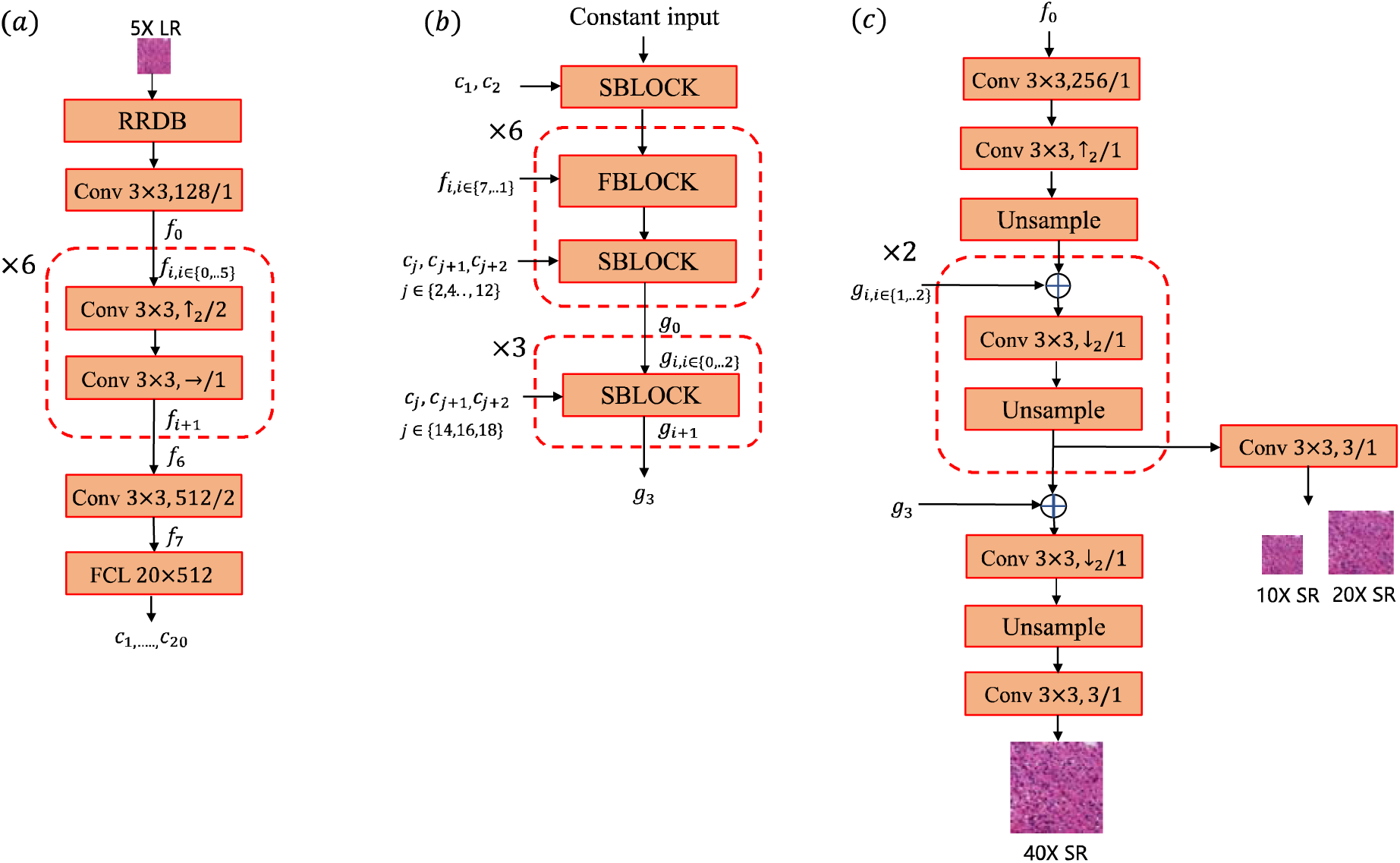
multi-scale SR model. (a) The RRDB-net [39] is employed for generating the latent vectors *c*_1→20_. Here, “conv” signifies a convolutional layer with attributes such as kernel size, number of output channels, and stride size. For simplicity, the subsequent LReLU activation function is omitted. ↑_2_ indicates a doubling of the number of channels, →indicates no change, and ↓_2_ denotes a reduction by half. *f, g* denote feature maps, and ×N denotes repetition *N* times. The FCL (fully connected layer) generates 20 latent vectors, each comprising 512 dimensions. (b) Latent vector *c* are fed into a SBLOCK module, i.e. style-transfer network [37]. The module integrates information from latent vectors, noise elements, and feature maps to generate 2X enlarged maps through upsampling. FBLOCK connects inputs through the smallest dimension and then undergoes two convolutional operations with conv 3 × 3, ↓_2_/1 , 3 × 3, 3/1. The value of *j* is 1 to 9. (c) The feature maps of the 5X image and the output g collaborate in decoding the SR images, utilizing a pixel shuffle operation for upsampling. The dissimilarity between the SR images at 10*X*-> 40X obtained through the decoder and the actual HR images is assessed using pixel loss, perceptual loss, and discriminator loss [40].The VGG16 network [41] is employed to compute image features, and the perceptual loss is computed by utilizing features extracted from the 21st layer. The discriminator is from the styleGAN architecture [38].

### MSR model training

Considering that the style transfer network is initially trained on natural images, it encounters difficulties in proficiently generating pathological images from latent vectors. As a result, the second module (depicted in Figure 2(b)) undergoes a pretraining process after FBLOCK is removed. This pretraining involves employing 40X patches from Dataset-SR-T adhering to the default hyperparameters of styleGAN[38]. Subsequently, the MSR model is trained, with the in the second module are initialized using pretrained styleGAN, while other modules are initialized randomly. The paired LR patches and HR patches in the training set from Dataset-SR-T are utilized for training which the validation set is employed to select hyperparameters that yield the highest PSNR and SSIM values. The selected hyperparameters are outlined in Supplemental Table 1. The code is built upon Python [43] and PyTorch [44], with the module construction utilizing the codebase from Openmmlab [45]. The implementation was carried out on a server with a Tesla A100 80GB GPU and 256 GB memory.

### SR WSIs generation

The 40X WSIs in Dataset-Ass and Dataset-CRC-Ass was segmented into non-overlapping patches of 2048×2048 dimensions, and down sampled to 5X patches, which was utilized to generate HR patches using our MSR model. The HR patches are arranged to form SR WSIs (Dataset-Ass-SR in Table 1 and Dataset-CRC-Ass-SR in Table 3).

### Diagnoses assessment settings

In order to accurately assess the diagnostic performance of SR WSIs, we employed a double-blind experimental design. First, the HR and SR WSIs from Dataset-Ass and Dataset-Ass-SR, were randomly divided into two groups. Each in two paired SR and HR WSI appeared in only one of the groups. Secondly, there pathologists were randomly assigned to diagnose one group first, and then the other group. To prevent mutual influence between the two groups, the multi-reader multi-case analysis (MRMC) was utilized. The time interval between the diagnoses of the two groups was set at one month. All diagnoses were based solely on the images themselves, with no additional clinical or examination information used other than the organ name. Therefore, in conducting a quantitative comparison between SR images and HR images for diagnostic purposes, the experiment controls for covariates that account for differences in sample complexity and pathologist diagnostic proficiency.

### The training and testing of CRC AI

From the Dataset-CRC (Table 2), The patches in randomly selected 80% WSIs were used for the training (Dataset-CRC-Training), while the patches in remaining 20 %WSIs were allocated for the test set (Dataset-CRC-Testing). Within the Dataset-CRC-Training, the patches in randomly selected 10% WSIs were further extracted to create the validation set. This entire process was repeated 10 times to train 10 versions of model-HR, facilitating the computation of statistical measures.

A vision transformer with the patch of 16×16 and input size of 224×224 were used to build a patch-level CRC model, named model-HR, implemented by TFViTForImageClassification library initialized from the model checkpoint at google/vit-base-patch16-224-in21k, which was then trained by the resized 224×224 patches from the Dataset-CRC-Training. The selected hyperparameters are listed in supplemented Table 2. We employed the clustering-based inference strategy described in reference [22] for patient-level testing on Dataset-CRC-Ass and Dataset-CRC-Ass-SR. If 2×2 continuous patches or cluster were identified as having cancer by the model-HR, the cancer may indeed exist on WSI. Clinically, multiple WSIs may be obtained for one patient. The inference on the patient level was based on positive sensitivity, that is, if all WSIs from the same patient were identified as negative (no cancer), then the patient was negative, otherwise the patient was positive.

### The finetune and testing of model-HR

In order to assess the fine-tuning on SR patches could enhance the model-HR’s performance on SR WSIs, we built the SR version of Dataset-CRC. Considering each patch with dimensions of 300×300 undergoes a 20X magnification, every 16 patches are randomly selected to compose an image with dimensions of 1200×1200. Subsequently, a central region measuring 1024×1024 is extracted from this image and down sampled, resulting in a 5X image with dimensions of 256×256. These processed images are then utilized as input for the proposed MSR model, producing 20X SR images with dimensions of 1024×1024. From the generated SR images, a central region of 900×900 is cropped and further divided into 20X SR patches, each measuring 300×300. Following the aforementioned steps, the Dataset-CRC-Training-SR is derived from Dataset-CRC-Training, while the Dataset-CRC-Testing-SR is derived from Dataset-CRC-Testing. Finally, the model-HR was fine-tuned on Dataset-CRC-Training-SR using the same hyperparameters as listed in Supplementary Table 1, except for setting the epoch to 50, and the model finetuned-SR was obtained. As model-HR was trained 10 times, there are consequently 10 versions of the corresponding finetuned-SR. The clustering-based inference strategy were employed for patient-level diagnosis [22], where if 3×3 continuous patches or cluster were identified as having cancer by the finetuned-SR, the optimal balance between sensitivity and specificity is achieved.

## Author Contributions

Conceptualization, G.Y., K.S.W.; Methodology, G.Y., K.S.W., K.S.; Investigation, G.Y., X.H.M., H.W.D.; Programming, M.X.Z, B.Q.B, M.Z.Z; Writing—original draft, G.Y., K.S.W., Y.N.Q, M.X.Z, and B.Q.B; Writing—review and editing, G.Y., H.W.D., K.S.W. and Y.H.G; Funding acquisition, G.Y., H.M.X., H.W.D., K.S.W and Y.H.G.; Resources, K.S.W., G.Y., and H.W.D.; Data curation, K.S.W., R.J.L., Y.S.C., and Y.W.; Supervision, G.Y. The author(s) read and approved the final manuscript.

### Fundings

This work was funded by grant supports from the National Natural Science Foundation of China (81972490 (K.S.W.)), and the Natural Science Foundation of Hunan Province (2023JJ30737 (G.Y.)), and the Tongxing Pathology Public Welfare Project from Peking Union Medical College Foundation (G.Y.), the Social Development Project of Science and Technology Department of Shaanxi Province (No. 2023-YBSF-465 (Y.H. G.)) and the Science and Technology Plan Project of Xi’an Science and Technology Bureau (No. 22YXYJ0108 (Y.H. G.)).

### Code Availability

The code for MSR training and image generation are available for download on GitHub at the following repository: https://github.com/CSU-BME/Pathology_SR.

### Data Availability

The training and validation datasets for the MSR model can be downloaded from GitHub at the following repository: https://github.com/CSU-BME/Pathology_SR. For access to the whole slide images in Dataset-Ass and Dataset-CRC for scientific research purposes, please contact the corresponding author.

---

**Supplementary Figure 1.**
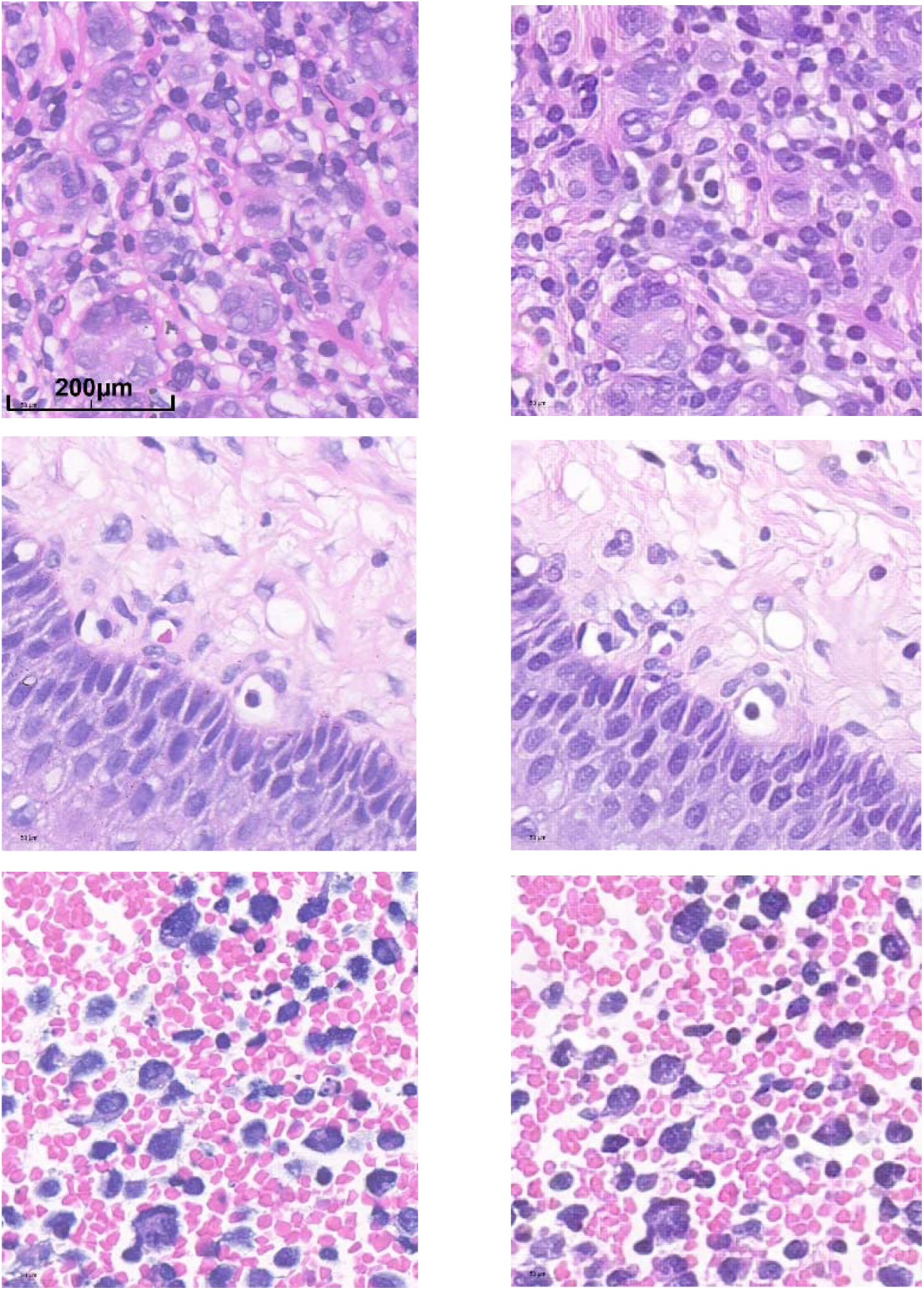
Visual comparison of some 40X HR and SR images. The left column consists of HR images, while the right column consists of SR images. For display, they are scaled to appropriate sizes.

**Supplementary Table 1.** List of hyperparameters for training MSR model Name of hyperparameters.

| **Name of hyperparameters** | **values** |
| --- | --- |
| Optimizer of generator | Adam |
| Optimizer of discriminator | Adam |
| betas | 0.9,0.99 |
| initial learning rate | 0.001 |
| final learning rate | 0.00001 |
| policy of learning rate | cosine restart |
| iteration number | 300000 |
| batch size | 2 |
| weight of MSE loss | 1.0 |
| weight of perceptual loss | 0.01 |
| weight of discriminator loss | 0.01 |

**Supplementary Table 2.** List of hyperparameters for AI model Name of hyperparameters.

| **Name of hyperparameters** | **values** |
| --- | --- |
| optimizer | Adam |
| epoch number | 500 |
| steps per epoch | 250 |
| batch size | 64 |
| learning rate | 0.0001 |
| decay rate | 0.99 |
| loss function | cross entropy |
| patience on early stopping | 50 |
| weight of L2 regulzrizer | 0.0001 |
| **Name of hyperparameters for finetune** | **values** |
| epoch number | 50 |
| steps per epoch | 250 |
| patience on early stopping | 20 |

